# Emergence of neutralizing antibodies associates with clearance of SARS-CoV-2 during HIV-mediated immunosuppression

**DOI:** 10.1101/2023.08.18.23293746

**Authors:** Farina Karim, Mallory Bernstein, Zesuliwe Jule, Gila Lustig, Janine-Lee Upton, Yashica Ganga, Khadija Khan, Kajal Reedoy, Matilda Mazibuko, Katya Govender, Kershnee Thambu, Nokuthula Ngcobo, Elizabeth Venter, Zanele Makhado, Willem Hanekom, Anne von Gottberg, Quarraisha Abdool Karim, Salim S. Abdool Karim, Nithendra Manickchund, Nombulelo Magula, Bernadett I. Gosnell, Penny L. Moore, Richard J. Lessells, Tulio de Oliveira, Mahomed-Yunus S. Moosa, Alex Sigal

## Abstract

To design effective vaccines and other immune interventions against a pathogen, it is necessary to know which aspect of immunity associates with protection. We investigated whether neutralizing antibodies associate with infection clearance in long-term SARS-CoV-2 infection during HIV-mediated immunosuppression. We monitored neutralizing antibody activity against SARS-CoV-2 over 1 to 2 years in five participants with advanced HIV disease and delayed control of HIV viremia. These participants had persistent SARS-CoV-2 infection ranging from 110 to 289 days which was associated with low or undetectable neutralizing antibody responses. SARS-CoV-2 clearance was associated with the emergence of neutralizing antibodies and occurred in two participants before suppression of HIV viremia, but after some CD4 T cell reconstitution. Vaccination only further increased neutralizing antibody levels in the advanced HIV disease participants who achieved HIV suppression pre-vaccination. During the prolonged SARS-CoV-2 infection we observed widespread evolution which was particularly pronounced in one Delta variant infection. This resulted in high-level escape from Delta-elicited neutralizing antibodies and a virus antigenically distinct from both ancestral SARS-CoV-2 and Omicron XBB in hamster experimental infections. The results offer new evidence that neutralizing antibodies associate with SARS-CoV-2 clearance and argue that successful management of HIV may be necessary to curtail long-term infection and evolution of co-infecting pathogens.

## Introduction

Neutralizing antibodies are a key correlate of protection against most viral infections^1,2^. During the Covid-19 pandemic, evolution of new viral variants reduced the ability of neutralizing antibodies elicited by previous infection or vaccination to prevent new symptomatic infections, although protection from more severe disease was generally maintained^3^. One possibility of how SARS-CoV-2 variants arise is evolution during long-term infection in immunosuppressed individuals^4–16^. Such evolution in immunocompromised hosts is not unique to SARS-CoV-2 and has been observed in influenza and salmonella infections^17,18^.

Case studies of SARS-CoV-2 infections in immunosuppressed individuals show prolonged infection and evolution of genomic changes in the SARS-CoV-2 spike protein associated with escape from neutralizing antibodies^13^. Mutations outside of spike are also common and may confer different functions^19,20^. SARS-CoV-2 long-term infection and evolution happens in some individuals who are immunosuppressed because of long-term uncontrolled HIV infection, termed advanced HIV disease^4–6,15^. Here, immunosuppression occurs because HIV infection depletes CD4 T cells by a variety of mechanisms that include death of both HIV-infected^21–24^ and uninfected bystander or incompletely infected cells^25–29^. Advanced HIV disease is defined as a CD4 T cell count lower than 200 cells/microliter (a normal CD4 T cell count is about 1000 cells/microliter). This level of CD4 T cell depletion is known to result in vulnerability to multiple pathogens. One example is *Mycobacterium tuberculosis*, one of the cardinal infections leading to the death of people living with HIV (PLWH) in the pre-antiretroviral therapy (ART) era^30,31^.

While there can be multiple reasons for immunosuppression^7,16,32–34^, the number of people globally with immunosuppression because of advanced HIV disease may be considerable. In South Africa alone, the estimated number of PLWH is about 8 million^35^. About 1 in 10 are thought to meet the criteria for advanced HIV disease^36,37^ - an estimated 800,000 people. Vaccination could potentially be a strategy to elicit a better immune response in people with advanced HIV disease. However, for SARS-CoV-2, vaccines were reported to be less effective at eliciting a neutralizing antibody response in PLWH with CD4 counts lower than 200 cells/microliter^38–40^.

We have previously reported one case where advanced HIV disease interferes with SARS-CoV-2 clearance and leads to SARS-CoV-2 evolution (participant 27 in this study)^4,5^. The virus which evolved in this participant over 6 months gained immune escape from neutralizing antibodies elicited by SARS-CoV-2 infection and Pfizer BNT162b2 mRNA vaccination. Here we tracked SARS-CoV-2 infection in five participants with advanced HIV disease and failure to adhere to ART, who eventually suppressed HIV viremia. All had prolonged SARS-CoV-2 infection. Clearance was associated with emergence of neutralizing antibodies. Among the viruses we isolated from these participants, SARS-CoV-2 originating in a Delta variant infection evolved the most antibody escape mutations and had high-level escape from Delta-elicited neutralizing antibodies. However, it did not escape the current population neutralizing antibody immunity to SARS-CoV-2.

## Results

### Advanced HIV disease leads to long-term SARS-CoV-2 infection and evolution

Five participants with advanced HIV disease and HIV viremia are described in this study. They are part of an observational longitudinal cohort of SARS-CoV-2 infection in South Africa numbering 994 participants, including 113 PLWH with a CD4 T cell count lower than 200 cells/microliter at enrollment (Table S1). Participant information was de-identified and participants assigned numbers. The linkage between number and identifying information is only known to the clinical team of the study. The five participants, whose de-identified numbers were 27, 96, 127, 255, and 209, were between 20 and 50 years old and had a Covid-19 diagnosis date ranging from September 2020 to December 2021 (Table S2). All participants were living with HIV before SARS-CoV-2 infection and participant 255 was HIV-infected by mother-to-child transmission. Participants were outpatients for 82% of study visits. During each study visit, a combined nasopharyngeal and oropharyngeal swab was taken to detect SARS-CoV-2 by qPCR cycle threshold (Ct, Figure 1A) which is inversely proportional to SARS-CoV-2 titer. The virus was isolated from the swab by outgrowth and/or sequenced when viral titers were sufficient (Ct<30 for sequencing and Ct<25 for isolation). The timepoints and titers of successfully sequenced samples are shown in Figure 1A as red circles. The duration of SARS-CoV-2 infection, calculated as time from first diagnostic to last qPCR positive SARS-CoV-2 test was a median of 207 days, and ranged from 110 to 289 days (Table S2). Four of the participants were enrolled soon after diagnosis. One participant (255) was enrolled in the study in December 2021 during the Omicron infection wave. However, a record of a positive qPCR result for SARS-CoV-2 was present from September 2021, corresponding the Delta variant infection wave (Figure 1A).

**Figure 1:**
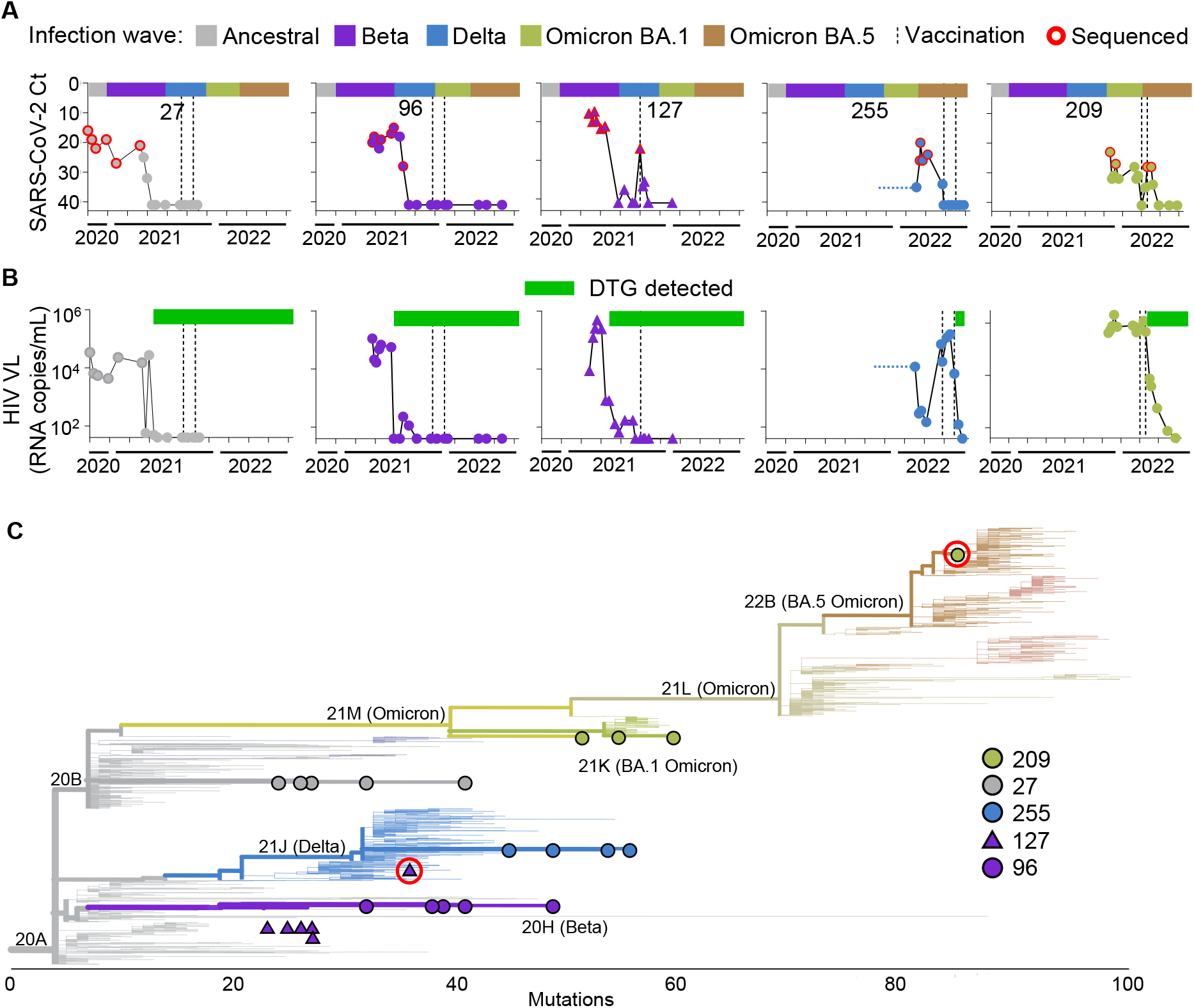
Persistent SARS-CoV-2 infection and accumulation of mutations in advanced HIV disease immunosuppression. A) SARS-CoV-2 infection through time in five participants with advanced HIV disease. X-axis represents infection period and bar above each graph represents the timing of the infection waves for each variant/strain in South Africa. Y-axis represents the qPCR cycle threshold (Ct) value, inversely proportional to the SARS-CoV-2 viral titer, as detected in the combined nasopharyngeal and oropharyngeal swab. Red circles represent successfully sequenced timepoints and vertical dashed lines represent Pfizer BNT162b2 mRNA vaccination times. B) HIV viral loads for the participants measured in the blood as RNA copies/mL. Green bars above graphs denote periods of adherence to dolutegravir (DTG) based ART. C) Phylogenetic tree of the sequenced virus samples through time for each participant (27: grey circles, 96: purple circles, 127: purple triangles, 127: blue circles, 209: green circles). X-axis represents distance in mutations from ancestral clade 20A SARS-CoV-2. Tree generated using Nextclade (https://clades.nextstrain.org/).

Participants were initiated on ART in line with current national guidelines and received adherence counselling from the clinical team. The ART regimen used was the integrase inhibitor dolutegravir (DTG), combined with the nucleotide/nucleoside reverse transcriptase inhibitors tenofovir (TFV) and lamivudine (3TC). The five participants described here had delayed control of HIV viremia (Figure 1B). Retrospective detection of ART levels in this group by liquid chromatography coupled with tandem mass spectrometry showed non-adherence to the DTG based regimen. Green bars in Figure 1B show the time when DTG started to be consistently detected and Figure S1 shows study visits where DTG, TFV, and 3TC were detected. Surprisingly, other antiretroviral drugs were also detected, possibly previously initiated regimens (Figure S1, blue rectangles). The participants eventually adhered to DTG based ART (Figure S1, green rectangles), leading to HIV suppression observed as a decline in HIV viremia to below the threshold of assay detection (40 HIV RNA copies/mL) during the study (Figure 1B).

Four of the participants were vaccinated with two doses of the Pfizer BNT162b2 mRNA vaccine and vaccination times are shown as vertical dashed lines in Figure 1A-B. The fifth (127) was vaccinated with only one dose because the participant developed synovial inflammation in the wrists and hands 6 days post-first vaccine dose, a rare adverse event associated with Covid-19 mRNA vaccines^41,42^. The interval between doses approximately followed the South African guidelines at the time of vaccination, which was 6 weeks, although some variation occurred.

We next determined the infecting variant by sequencing. Alignment of sequenced SARS-CoV-2 showed that, consistent with the infection date (Figure 1A), the participant designated 27 was infected with ancestral SARS-CoV-2 (Figure 1C, grey circles) and the participant designated 96 was infected with the Beta variant (Figure 1C, purple circles). Participant 127 was initially infected with a Beta variant (Figure 1C, purple triangles) but the last sequence was a Delta variant (Figure 1C, highlighted in red), likely a re-infection which occurred during the Delta infection wave in South Africa. Participant 255 was infected with the Delta variant (Figure 1C, blue circles), consistent with a continuous infection from the first positive diagnostic test in September 2021. Participant 209 was infected with the Omicron BA.1 subvariant (Figure 1C, green circles), but the sequence from the last timepoint was an Omicron BA.5 subvariant (Figure 1C, highlighted in red), likely a re-infection.

We next analyzed non-synonymous changes across the SARS-CoV-2 genome from each sequenced timepoint per participant using the Stanford Coronavirus Antiviral and Resistance Database (https://covdb.stanford.edu/sierra/sars2/by-sequences/). Three of the participants, 27, 96 and 255, had extensive non-synonymous changes in the circulating virus, while two, 127 and 209, had few changes but showed an abrupt shift in sequence consistent with re-infection with a different variant or sub-variant (Figure S2). Both 27 and 255 had multiple substitutions in the receptor binding domain (RBD) of the spike gene predicted to lead to escape from neutralizing antibodies. These included E484K, K417T and F490S^43–47^ in the virus from participant 27 and K417T, L452Q, A475V, and E484A^46–50^ in the virus from participant 255.

### Long-term SARS-CoV-2 infection clearance associates with emergence of neutralizing antibodies

To investigate the relationship between the neutralizing antibody response and SARS-CoV-2 clearance, we isolated and expanded at least one SARS-CoV-2 virus from each participant and tested the neutralizing capacity of the participant’s plasma against the autologous outgrown viruses at the different timepoints post-diagnosis. Figure 2A shows SARS-CoV-2 viral titers through time for each participant up to and including the virus clearance timepoint, where clearance was defined as two or more consecutive timepoints where SARS-CoV-2 is undetectable (see Figure 1A).

**Figure 2:**
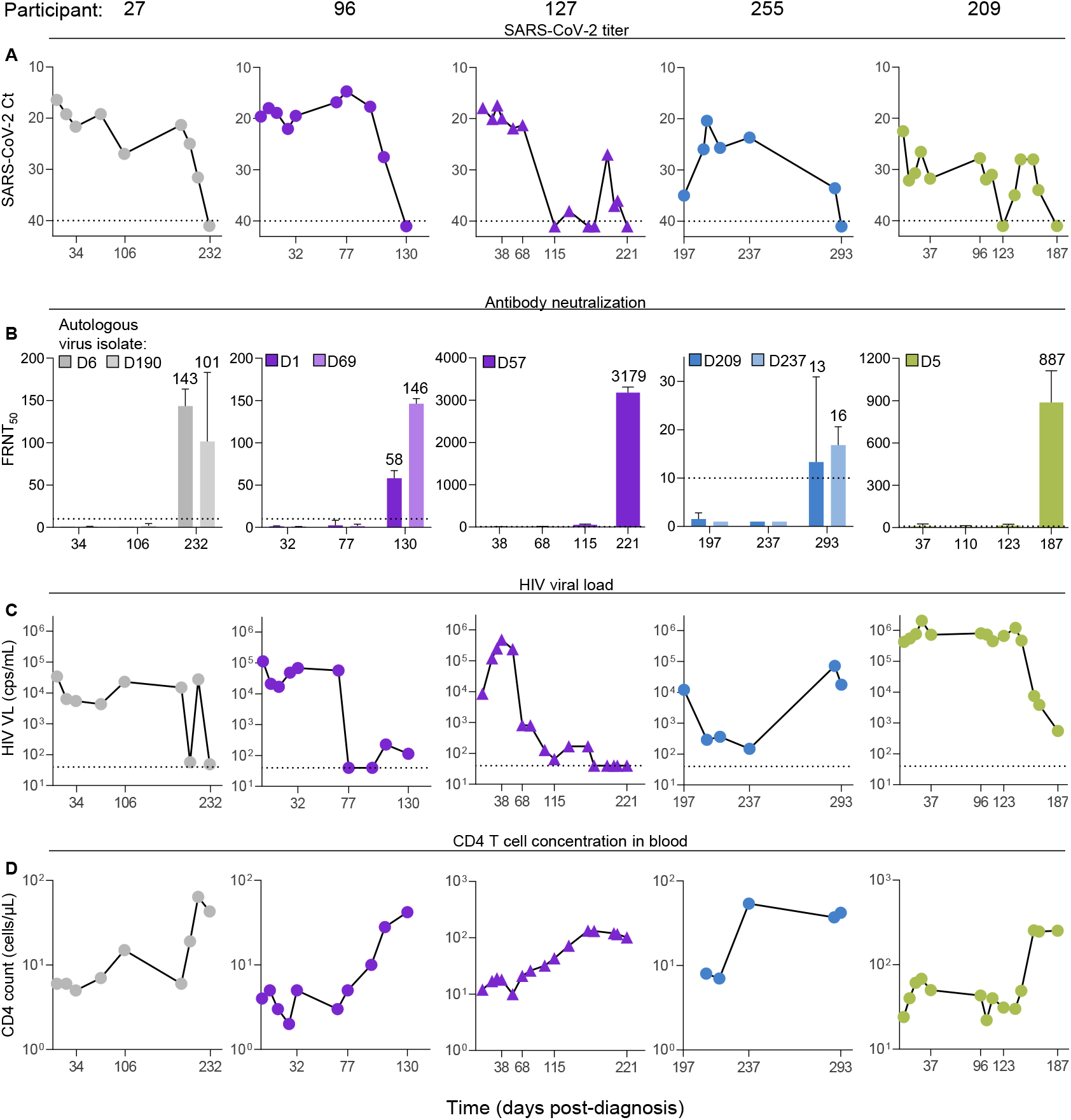
SARS-CoV-2 clearance in advanced HIV disease immunosuppression associates with emergence of a neutralizing antibody response. A) SARS-CoV-2 titers measured as qPCR Ct through time until SARS-CoV-2 clearance for participants 27, 96, 127, 255, and 209. Clearance defined as two consecutive timepoints where SARS-CoV-2 was not detected. X-axis is time in days post-SARS-CoV-2 diagnosis and is the same in all panels which describe the same participant. Y-axis is SARS-CoV-2 titer as Ct value. B) Antibody neutralization of SARS-CoV-2 isolated from each participant. One to two autologous viruses were tested per participant and are indicated on the top left of each graph by the day of isolation (denoted by a D prefix) post-diagnosis. Numbers above bars are geometric means and error bars indicate geometric mean standard deviations of 2-4 independent experiments. C) HIV viral load in the blood during the period of SARS-CoV-2 infection. D) CD4 T cell concentrations during the period of SARS-CoV-2 infection. Horizontal lines represent limits of quantification in all panels.

The neutralization capacity of participant plasma was determined throughout this work by a focus forming assay with the live virus isolates (Figure S3). To quantify the result, we present the focus reduction neutralization test 50 value (FRNT_50_), the inverse of the plasma dilution required for 50% neutralization. Ongoing SARS-CoV-2 infection was correlated with low FRNT_50_ values at or below the level of detection (Figure 2B). Strikingly, in all cases, there was a strong increase in neutralization capacity against the autologous viruses at SARS-CoV-2 clearance (Figure 2B, last timepoint in each graph). The absolute FRNT_50_ value which was associated with viral clearance varied between participants. In participant 96 it also varied between virus isolates. In participant 255, who had undetectable neutralizing antibody levels before SARS-CoV-2 clearance, the FRNT_50_ value became detectable but remained relatively low.

We examined the HIV viral load (Figure 2C) and CD4 T cell counts (Figure 2D) at each timepoint per participant. While participants 27, 96, and 127 showed HIV suppression at SARS-CoV-2 clearance, participant 209 did not have full suppression (Figure 2C, rightmost panel) and participant 255 was unsuppressed, with an HIV viral load of about 10^4^ RNA copies/mL (Figure 2C, panel second from right). Immune reconstitution measured in terms of an increase in CD4 T cell counts occurred in every participant (Figure 2D), although CD4 counts remained at 100 cells/microliter or below for 4 out of 5 participants. Participants had a gradual increase in CD4 count to the time of SARS-CoV-2 clearance. However, participant 255, who did not have a suppressed HIV viral load at clearance, showed CD4 T cell reconstitution which plateaued at a timepoint where SARS-CoV-2 was not yet cleared (Figure 2D, second from right).

### Poor vaccine neutralizing antibody response in advanced HIV disease participants with HIV viremia

We investigated the neutralizing antibody response elicited by vaccination in the advanced HIV disease participants and compared the response to participants, either PLWH or HIV negative, who did not have immunosuppression (Tables S3-S4). All participants received the Pfizer BNT162b2 mRNA vaccine.

We tested for neutralizing antibodies against ancestral virus, the Beta, Delta and Omicron BA.1 variants, as well as anti-spike antibody levels, at baseline and after each vaccine dose (Figure 3A). In the three participants with suppressed HIV at vaccination (27, 96, and 127), there was an increase in neutralization capacity for all strains/variants tested after the first, and if administered, second dose of the vaccine, and a similar increase in overall anti-spike antibodies. In participant 127, who received only one vaccine dose, vaccine elicited neutralization waned quickly, with neutralization capacity dropping approximately 10-fold in about 4 months against all viruses tested. In participant 27, neutralizing antibodies to Omicron BA.1 showed rapid waning (Figure 3A).

**Figure 3:**
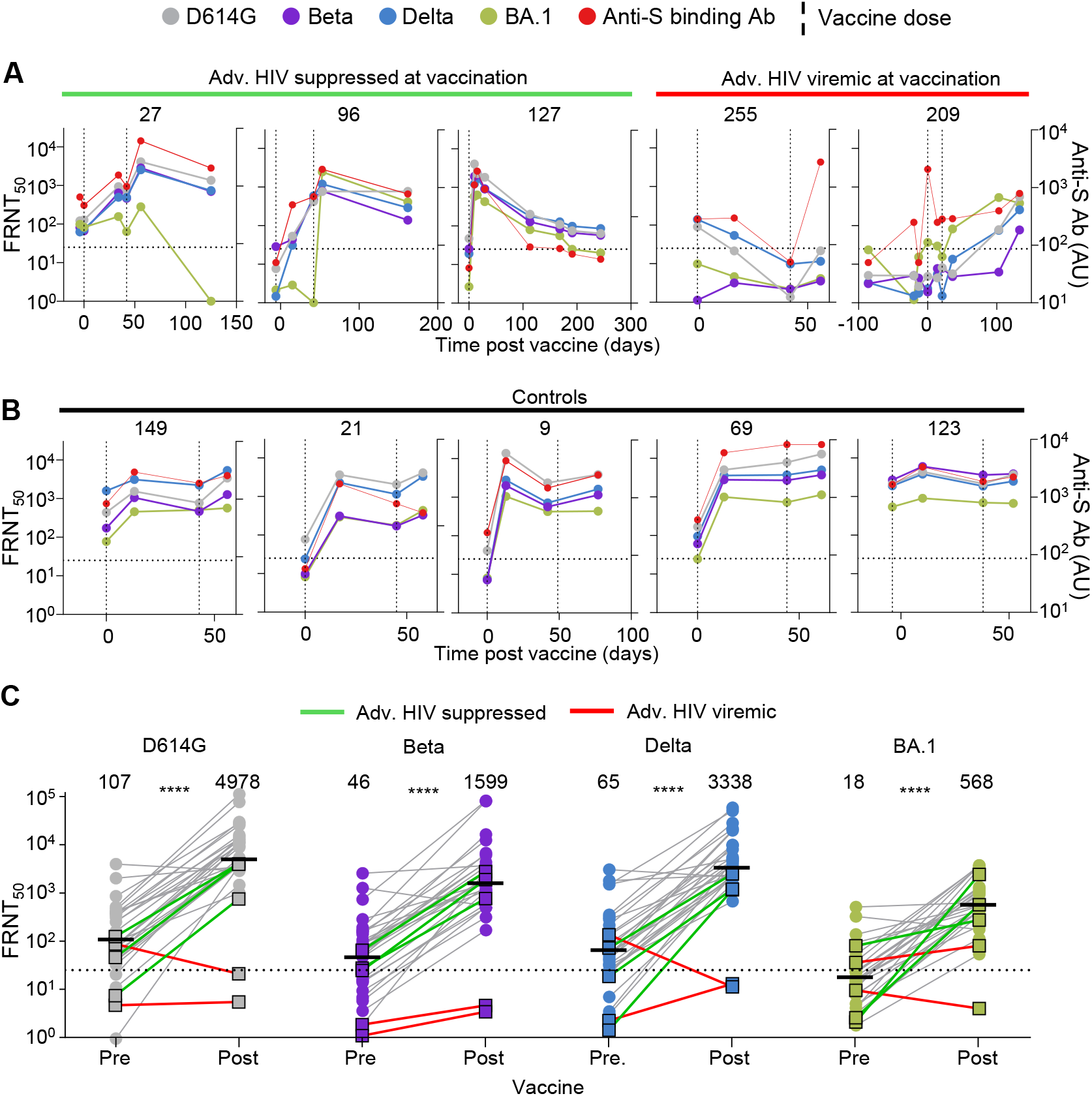
Advanced HIV disease participants with unsuppressed viremia have a poor vaccine response. A) Longitudinal neutralization and anti-spike antibody levels before and after vaccination with the Pfizer BNT162b2 mRNA vaccine in the three advanced HIV disease participants who suppressed HIV at vaccination (27, 96, 127) and the two who did not (255, 209). Neutralization was tested against ancestral/D614G SARS-CoV-2 (grey), Beta variant (purple), Delta variant (blue), and Omicron BA.1 subvariant (green). Timing of vaccine doses is represented by vertical dashed lines. X-axis is time in days post-first vaccine dose and negative numbers represent pre-vaccine period. Left Y-axis is neutralization capacity per viral isolate as FRNT_50_ and right Y-axis is anti-spike antibody level as arbitrary units (AU). B) Longitudinal neutralization and anti-spike antibody levels before and after vaccination in five participants with no advanced HIV disease. C) Neutralization of SARS-CoV-2 D614G, Beta, Delta and Omicron BA.1 viral isolates pre-vaccination and after last administered dose by plasma from the two participants with advanced HIV disease and HIV viremia (marked with red lines), the three participants with advanced HIV disease and HIV suppression (green lines) and 26 participants with no advanced HIV disease (grey lines). Y-axis is neutralization as FRNT_50_. Numbers above groups are geometric means and statistical comparison is between participant FRNT_50_ values before and after vaccination and are all ****p<0.0001 by the Mann-Whitney test.

In participants 255 and 209, HIV viremia was present at vaccination (Figure 1B). Participant 255 cleared SARS-CoV-2 at the vaccine baseline visit, before vaccination was administered. Participant 255 still had SARS-CoV-2 infection at vaccination (Figure 1A). Both 255 and 209 showed a poor initial response to the vaccine (Figure 3A). For 255, neutralization capacity decreased to below limit of quantification after the first dose and remained at that level after the second dose. For 209, neutralization capacity increased with time, but not at two weeks post-vaccination, the first post-vaccination timepoint tested and expected peak of the neutralization response to the vaccine (Figure 3A, right panel).

To compare our results to participants without advanced HIV disease who were also previously SARS-CoV-2 infected, we tested the antibody neutralization response after each vaccine dose in five participants with either suppressed HIV or who were HIV negative (see Table S3 for participant details). These participants showed a relatively homogenous response, with a large fold-increase post-first dose, usually followed by limited waning, then a smaller fold-increase in neutralizing capacity post-second dose (Figure 3B).

Interestingly, we observed that anti-spike binding antibody levels mirrored well the neutralizing antibody response to the different viral isolates. The exceptions were the two advanced HIV disease participants with HIV viremia at vaccination. For participant 255, the first timepoint post-vaccination showed a strong increase in anti-spike antibodies but not neutralization capacity, and this was also observed for participant 209 (who also had uncleared SARS-CoV-2) at the baseline visit before administration of the first antibody dose.

We quantified the response post-second dose in a larger group of previously infected participants with no advanced HIV disease (Table S4), who were PLWH (n=10) or HIV negative (n=16). This group included the control group of five participants with detailed longitudinal samples described above. In all participants, we tested for neutralizing antibodies against ancestral virus, the Beta, Delta and Omicron BA.1 variants (Figure 3C). The three advanced HIV disease participants with ART suppressed HIV at the time of vaccination showed a vaccine mediated increase in neutralization of all four viruses (Figure 3C, green lines). In contrast, neutralizing antibody remained low against the four viruses in the two advanced HIV disease participants with unsuppressed HIV (Figure 3C, red lines). Unlike the responses of the advanced HIV disease participants with suppressed HIV, these responses were distinct from the neutralizing antibody vaccine responses of the participants with no advanced HIV disease.

### Low XBB.1.5-infection elicited cross-neutralization of ancestral and Delta-evolved viruses in hamster infections

The degree to which neutralizing antibody immunity elicited by infection with one virus strain can cross-neutralize a second strain is a measure of the antigenic distance between them. However, given that SARS-CoV-2 seroprevalence in South Africa was approximately 70% pre-Omicron^51^, we could not use human sera to measure antigenic distance between recent Omicron subvariants such as XBB and other strains as we would be unlikely to find XBB infected individuals who were uninfected with earlier variants. We have therefore investigated cross-neutralization of ancestral SARS-CoV-2, the Omicron XBB.1.5 subvariant, and two of the evolved SARS-CoV-2 strains with the most antibody escape mutations (Figure 4A) in the Syrian golden hamster experimental infection model. The two evolved viruses were the virus evolved from ancestral SARS-CoV-2 in participant 27 after a 190-day infection (27-D190), and the virus evolved from the Delta variant in participant 255 after a 237-day infection (255-D237). Sixteen days post-infection, plasma samples from uninfected and infected animals were tested against the autologous (infecting) virus as well as the three other viruses to determine cross-neutralization (Figure 4B).

**Figure 4:**
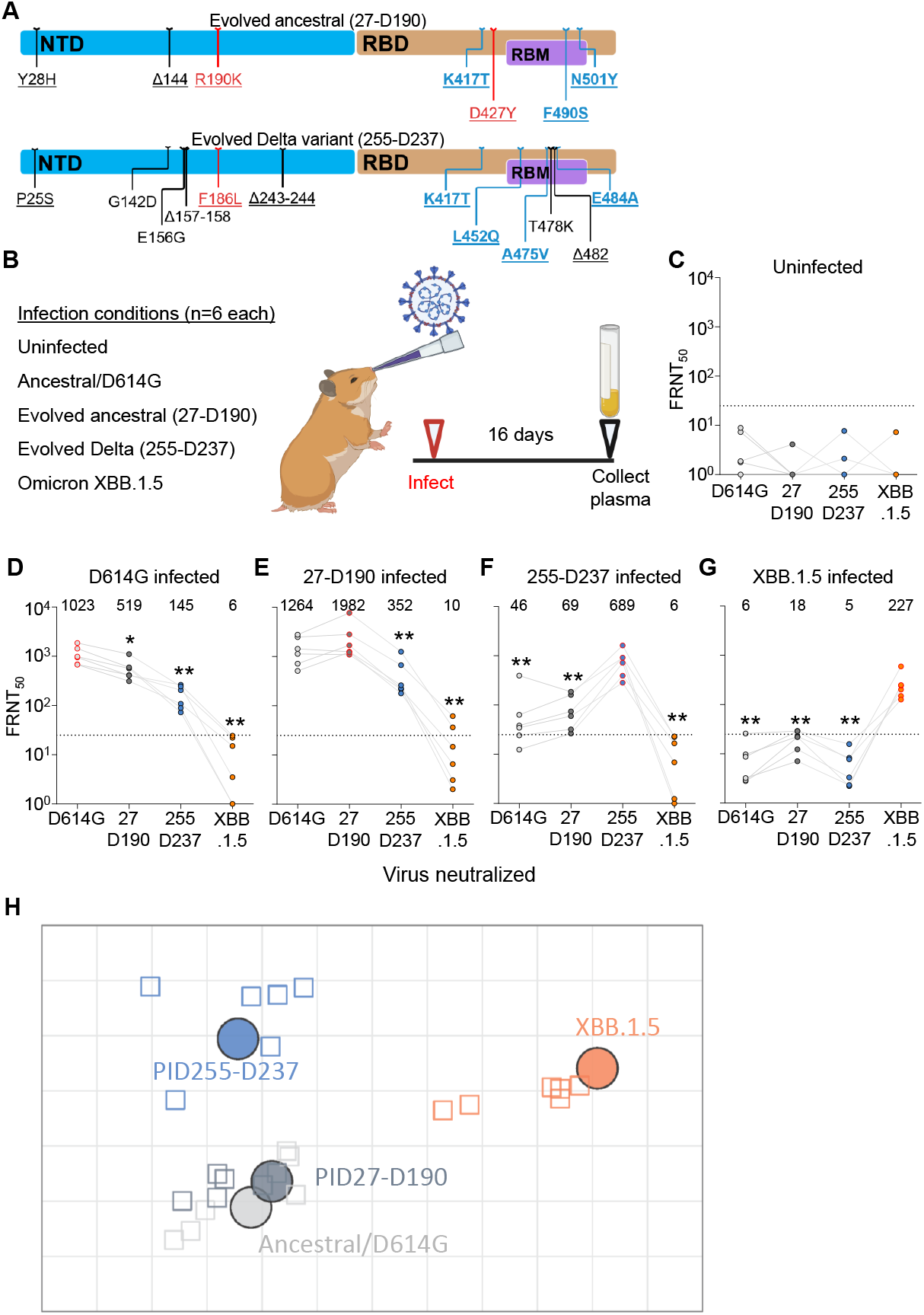
Antigenic distances of SARS-CoV-2 evolved from ancestral and Delta infections in the hamster model. A) Substitutions and deletions in the N-terminal domain (NTD) and receptor binding domain (RBD) of SARS-CoV-2 spike in 27-D190 and 255-D237. Blue mutations: known antibody escape. Red mutations: mutations with global prevalence below 0.01%. RBM: receptor binding motif. Representation and characterization based on the Stanford Coronavirus Antiviral and Resistance Database (https://covdb.stanford.edu). B) Schematic of hamster infection experiment. Six animals in two independent experiments were used per infection condition. C) Neutralization of ancestral D614G, 27-D190, 255-D237, and Omicron XBB.1.5 subvariant viruses in uninfected hamsters. D-G) Neutralization of the same viruses at 16 days post-infection in hamsters infected with ancestral/D614G (D), 27-D190 (E), 255-D237 (F), and the XBB.1.5 subvariant (G). Numbers above the points and horizontal bars are geometric means for the group. Significance was determined by the Mann-Whitney test relative to the autologous (infecting) virus. Significant p-values from left to right were: 0.03, 0.002, 0.002. 0.009, 0.002. 0.009, 0.002, 0.002, 0.002, 0.002, 0.002. H) Antigenic map of neutralization data presented in panels D-G. Virus strains/variants are shown as colored circles and hamster plasma samples as open squares with the color corresponding to the virus strain or variant used to infect the hamster. Each square on the grid correspond to a twofold decrease in neutralizing capacity. Map created using Racmacs (https://acorg.github.io/Racmacs/). Parts of panel B created with BioRender.com.

Plasma from uninfected animals did not neutralize any of the viruses (Figure 4C). Plasma from infected animals most potently neutralized the autologous virus and cross-neutralized the other viruses less well (Figure 4D-G). The animals not infected with Omicron XBB.1.5 did not have a substantial cross-neutralization of XBB.1.5 (Figure 4D-F), and animals infected with XBB.1.5 failed to develop a cross-neutralizing response to the other viruses tested (Figure 4G).

To better visualize the antigenic distances between the viruses, we used antigenic cartography (Figure 4H) which maps the distances between multiple viruses and the sera elicited by their infections^52–54^. Here, each square of distance corresponds to a 2-fold drop in neutralizing capacity. Using this visualization, we observed that 27-D190 was antigenically close to ancestral virus from which it evolved. In contrast, XBB.1.5 was antigenically far from all the other viruses tested. The Delta evolved 255-D237 was antigenically distinct from both ancestral SARS-CoV-2 and Omicron XBB.1.5.

### Evolved Delta variant escapes Delta but not Omicron XBB-elicited neutralizing antibodies

People were likely infected through the course of the pandemic with multiple SARS-CoV-2 strains/variants, and therefore their neutralizing antibody responses may have a different pattern than those we observed in the singly infected hamsters. As we have previously characterized the 27-D190 evolved virus^4,20^, here we tested the Delta variant evolved 255-D237 virus which had mutations predicted to result in immune escape from neutralizing antibodies (Figure 4A). We tested this virus against plasma panels from participants infected in the Delta, Omicron subvariant BA.1, and Omicron subvariant XBB infection periods in South Africa (see Table S5 for participant details).

We first tested 255-D237 against plasma from participants infected with the Delta variant. We observed that, relative to the Delta variant virus, the 255-D237 had over 18-fold lower FRNT_50_ (Figure 5A), escape similar to Omicron BA.1 relative ancestral virus in participants vaccinated with ancestral virus based vaccines^55^. We then tested this virus against plasma from Omicron BA.1 infected participants. As we reported previously, we found Omicron BA.1 elicited neutralizing immunity to be relatively low^56^. The 255-D237 evolved virus had a 6.5-fold escape of neutralization relative to the Omicron BA.1 virus in this group, with some of the neutralization values for 255-D237 falling below the threshold of quantification (Figure 5B).

**Figure 5:**
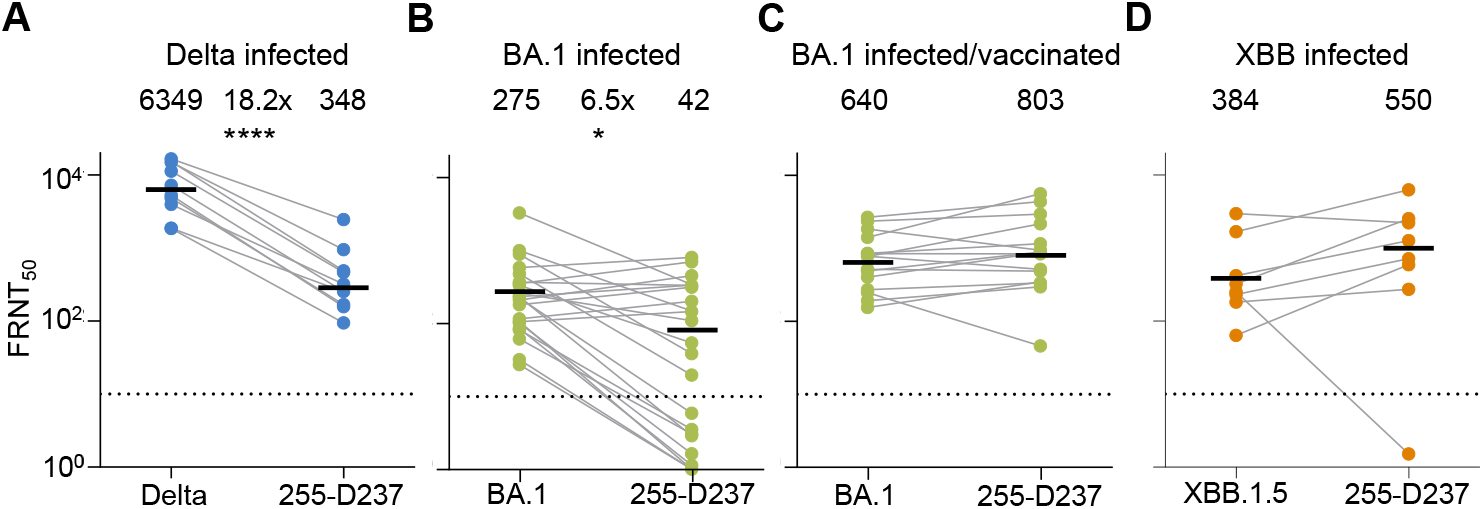
Neutralization of Delta evolved virus by participants with Delta and Omicron infection elicited immunity. A) Neutralization of the 255-D237 virus compared to the Delta variant virus by plasma samples from 10 participants infected during the Delta variant infection wave in South Africa. B) Neutralization of the 255-D237 virus compared to the Omicron BA.1 subvariant virus in plasma samples from 24 participants infected with Omicron BA.1. FRNT_50_ for one participant was out of the range of the graph but included in the calculation of the geometric mean. C) Neutralization of 255-D237 compared to Omicron BA.1 virus in plasma samples from 15 vaccinated participants with Omicron BA.1 breakthrough infection. D) Neutralization of the 255-D237 compared to the Omicron XBB.1.5 subvariant virus in plasma samples from 8 participants infected when XBB subvariants were dominant in South Africa. All samples were collected approximately 2 to 3 weeks post-diagnosis in the corresponding infection period. The numbers above the points and horizontal bars are geometric means of the group and p-values were determined by the Mann-Whitney test and were: ****p<0.0001, *p=0.03.

We then proceeded to test 255-D237 against plasma from vaccinated participants with breakthrough Omicron BA.1 infection. As we previously reported^56^, the vaccinated/BA.1 infected group had stronger Omicron BA.1 virus neutralization as well as better cross-neutralization of other variants compared to the unvaccinated group. We observed that neutralizing antibody immunity in this group neutralized the 255-D237 virus to a similar extent as the BA.1 virus (Figure 5C). Lastly, we tested against plasma from individuals infected during the period when Omicron XBB subvariants were dominant in South Africa (from November-December 2022, https://www.nicd.ac.za/diseases-a-z-index/disease-index-covid-19/sars-cov-2-genomic-surveillance-update/ accessed August 8, 2023). These participants are also expected to have immunity from previous infections, including pre-Omicron variants. In this group there was no escape of the 255-D237 virus relative to the XBB.1.5 subvariant (Figure 5D).

## Discussion

Here we characterized five long-term SARS-CoV-2 infections in individuals with advanced HIV disease who had HIV viremia upon study enrollment. We found that SARS-CoV-2 clearance was associated with the emergence of a neutralizing antibody response in every case. Based on vaccine trials, neutralizing antibodies are a known correlate of protection against symptomatic SARS-CoV-2 infection^2^. However, this study is, to our knowledge, the first to demonstrate an association between the emergence of an antibody response and SARS-CoV-2 clearance in long-term infection in a group of participants. The study has the limitation that, despite the large number of participants followed in our cohort, the number of participants with advanced HIV disease and persistent HIV viremia was small. The potentially key role of the neutralizing antibody response in clearing SARS-CoV-2 which we found in this study is consistent with observations that B cell depletion with rituximab leads to persistent SARS-CoV-2 infection^57–60^.

We observed that the neutralizing antibody response did not need to be very strong to associate with SARS-CoV-2 clearance, as demonstrated for participant 255. A related observation was that SARS-CoV-2 was cleared despite HIV viremia being present. Incomplete HIV suppression at SARS-CoV-2 clearance was also observed in participant 209. Some reconstitution of CD4 T cell numbers occurred in all participants, including the HIV viremic participants, prior to SARS-CoV-2 clearance. CD4 T cell reconstitution likely occurred because of ART initiation, even if initiation was partial. Since T cell help is essential to generate an effective antibody response^61^, this reconstitution may have led to the production of neutralizing antibodies to SARS-CoV-2.

We found that the Pfizer BNT162b2 mRNA vaccine was effective in increasing binding and neutralizing antibodies against SARS-CoV-2 variants in participants with advanced HIV disease who had ART suppressed HIV viremia. It was not effective in the two participants in whom HIV viremia was still present. This agrees with previous studies showing a less effective neutralizing antibody responses to vaccines in individuals with low CD4 T cell counts^38–40^. Therefore, vaccination in the absence of HIV suppression may not be an effective strategy to increase immunity in people with advanced HIV disease.

Interestingly, while binding antibody levels tracked closely with neutralization capacity, the two viremic advanced HIV disease participants showed instances where binding antibodies were elevated without a rise in neutralizing antibody activity, perhaps because of a lack of neutralization potency, which can be associated with poor B cell affinity maturation^62,63^.

All advanced HIV disease participants evolved SARS-CoV-2 mutations in their circulating viruses in long-term infection. Two of the participants, infected with ancestral and Delta variant viruses respectively, had evolved multiple known antibody escape mutations including E484K, E484A, K417T, F490S, L452Q, and A475V^43–50^. Examining the consequences of these mutations in the hamster infection model showed that the virus evolved from the Delta variant had considerable antigenic distance from both ancestral SARS-CoV-2 and the Omicron XBB.1.5 subvariant. Interestingly, hamster infection with XBB.1.5 failed to elicit neutralizing immunity not only to the Delta variant evolved virus but also to ancestral SARS-CoV-2, in agreement with recent results^64^. The Delta variant evolved virus also showed extensive escape from immunity elicited by Delta variant infections in people. However, it did not escape more recent neutralizing immunity from vaccinated individuals with Omicron BA.1 breakthrough infection, nor from neutralizing immunity elicited by Omicron XBB subvariants. It is likely that most of the BA.1 and XBB infected group have immunity from pre-Omicron infections^51,65^, and in the case of the XBB group, other Omicron subvariant infections^66^. Therefore, in contrast to the hamster experimental infections which used one virus, individuals enrolled during the XBB period likely had hybrid immunity from multiple exposures, leading to wider neutralization breadth^56^ and accounting for the difference in the results.

In conclusion, we have found new evidence that neutralizing antibodies associate with SARS-CoV-2 clearance and are likely required for such clearance to happen. However, the levels need not be high. Furthermore, we have found that, while SARS-CoV-2 can evolve extensively in long-term infection in advanced HIV disease, neutralizing antibody immunity from recent infections, most likely combined with that acquired in past infections, may neutralize some of the viruses evolved in this way. It is likely that such evolution during long-term infection in immunosuppressed individuals occurs in other pathogens in HIV co-infections. Therefore, investment in an effective global HIV treatment strategy may be necessary to reduce the chances that this type of evolutionary process results in a pathogen with pandemic potential.

## Methods

### Informed consent and ethical statement

All blood samples used for neutralization studies, nasopharyngeal swabs from the advanced HIV disease participants for outgrowth and sequencing, as well as nasopharyngeal swabs for isolation of the ancestral/D614G, Beta, and Delta virus were obtained after written informed consent from adults with PCR-confirmed SARS-CoV-2 infection enrolled in a prospective cohort of SARS-CoV-2 infected individuals at the Africa Health Research Institute approved by the Biomedical Research Ethics Committee at the University of KwaZulu–Natal (reference BREC/00001275/2020). The Omicron/BA.1 virus was isolated from residual swab used for diagnostic testing (Witwatersrand Human Research Ethics Committee reference M210752). The nasopharyngeal swab for isolation of the Omicron XBB.1.5 subvariant was collected after written informed consent as part of the COVID-19 transmission and natural history in KwaZulu-Natal, South Africa: Epidemiological Investigation to Guide Prevention and Clinical Care in the Centre for the AIDS Programme of Research in South Africa (CAPRISA) study and approved by the Biomedical Research Ethics Committee at the University of KwaZulu–Natal (reference BREC/00001195/2020, BREC/00003106/2021).

### Reagent availability statement

Viral isolates are available upon reasonable request. Sequences of isolated SARS-CoV-2 used in this study have been deposited in GISAID with accession numbers as follows:

| Virus | GISAID | Virus | GISAID |
| --- | --- | --- | --- |
| **D614G** | EPI_ISL_602626.1 | **0127-D10** | EPI_ISL_16508746 |
| **Beta** | EPI_ISL_678615 | **0127-D24** | EPI_ISL_16508747 |
| **Delta** | EPI_ISL_3118687 | **0127-D31** | EPI_ISL_16508748 |
| **BA.1** | EPI_ISL_7886688 | **0127-D38** | EPI_ISL_16508749 |
| **0027-D6** | EPI_ISL_15541746 | **0127-D54** | EPI_ISL_18030392 |
| **0027-D20** | EPI_ISL_15541747 | **0127-D68** | EPI_ISL_16508751 |
| **0027-D34** | EPI_ISL_15541748 | **0127-D192** | EPI_ISL_14666773 |
| **0027-D71** | EPI_ISL_15541749 | **0255-D209** | EPI_ISL_14599778 |
| **0027-D106** | EPI_ISL_15541750 | **0255-D211** | EPI_ISL_14599779 |
| **0027-D190** | EPI_ISL_2397313 | **0255-D219** | EPI_ISL_14599780 |
| **0096-D1** | EPI_ISL_14666761 | **0255-D237** | EPI_ISL_13986497 |
| **0096-D15** | EPI_ISL_14666763 | **0209-D5** | EPI_ISL_18030393 |
| **0096-D32** | EPI_ISL_13986492 | **0209-D26** | EPI_ISL_18030394 |
| **0096-D68** | EPI_ISL_18030390 | **0209-D144** | EPI_ISL_12970433 |
| **0096-D77** | EPI_ISL_18030391 | **0209-D159** | EPI_ISL_14666777 |
| **0096-D110** | EPI_ISL_14666766 |  |  |

### Clinical laboratory testing

SARS-CoV-2 Ct and HIV viral load quantification was performed from a nasopharyngeal swab universal transport medium aliquot and 4 ml EDTA tube of blood respectively at an accredited diagnostic laboratory (Molecular Diagnostic Services, Durban, South Africa). The CD4 count was performed by an accredited diagnostic laboratory (Ampath, Durban, South Africa).

### Detection of ART concentrations in plasma by LC-MS/MS

Sample analysis was performed using an Agilent High Pressure Liquid Chromatography (HPLC) system coupled to the AB Sciex 5500, triple quadrupole mass spectrometer equipped with an electrospray ionization (ESI) TurboIonSpray source. The LC-MS/MS method was developed and optimized for the quantitation of tenofovir, lamivudine and dolutegravir in the same sample. A protein precipitation extraction method using acetonitrile was used to process 50 μL plasma samples. 50 μL of water and 50 μL of ISTD solution was added and the sample was briefly mixed. 150 μL of acetonitrile was subsequently added to facilitate protein precipitation, vortex mixed and centrifuged at 16000 g for 10 min at 4°C. 170 μL of the clear supernatant was then transferred to a clean micro-centrifuge tube and dried down using a SpeedVac dryer set at 40°C. The dried samples were then reconstituted in 100 μL of 0.02% sodium deoxycholate (Sigma) in Millipore filtered water, vortex mixed, briefly centrifuged, placed in a small insert vial, capped, placed in the auto sampler compartment (maintained at 4°C) and analyzed using LC-MS/MS. The analytes were separated on an Agilent Zorbax Eclipse Plus C18 HPLC column using gradient elution. The column oven was set at 40°C, a sample volume of 2 μL was injected and the flow rate was set to 0.2 mL/min. Mobile phase A consisted of water with 0.1% formic acid and B consisted of acetonitrile with 0.1% formic acid. The drug analytes were monitored using multiple-reaction monitoring mode for positive ions except for efavirenz which was monitored in the negative ion scan mode. Analyst software, version 1.6.2 was used for quantitative data analysis. Blanked values for EFV, FTC and TFV were in the range of 3 ng/mL and this was set as the detection limit.

### Whole-genome sequencing

RNA was extracted on an automated Chemagic 360 instrument, using the CMG-1049 kit (Perkin Elmer, Hamburg, Germany). The RNA was stored at −80◦C prior to use. Libraries for whole genome sequencing were prepared using either the Oxford Nanopore Midnight protocol with Rapid Barcoding or the Illumina COVIDseq Assay. For the Illumina COVIDseq assay, the libraries were prepared according to the manufacturer’s protocol. Briefly, amplicons were tagmented, followed by indexing using the Nextera UD Indexes Set A. Sequencing libraries were pooled, normalized to 4 nM and denatured with 0.2 N sodium acetate. An 8 pM sample library was spiked with 1% PhiX (PhiX Control v3 adaptor-ligated library used as a control). We sequenced libraries on a 500-cycle v2 MiSeq Reagent Kit on the Illumina MiSeq instrument (Illumina). On the Illumina NextSeq 550 instrument, sequencing was performed using the Illumina COVIDSeq protocol (Illumina Inc, USA), an amplicon-based next-generation sequencing approach. The first strand synthesis was carried using random hexamers primers from Illumina and the synthesized cDNA underwent two separate multiplex PCR reactions. The pooled PCR amplified products were processed for tagmentation and adapter ligation using IDT for Illumina Nextera UD Indexes. Further enrichment and cleanup was performed as per protocols provided by the manufacturer (Illumina Inc). Pooled samples were quantified using Qubit 3.0 or 4.0 fluorometer (Invitrogen Inc.) using the Qubit dsDNA High Sensitivity assay according to manufacturer’s instructions. The fragment sizes were analyzed using TapeStation 4200 (Invitrogen). The pooled libraries were further normalized to 4nM concentration and 25 μL of each normalized pool containing unique index adapter sets were combined in a new tube. The final library pool was denatured and neutralized with 0.2N sodium hydroxide and 200 mM Tris-HCL (pH7), respectively. 1.5 pM sample library was spiked with 2% PhiX. Libraries were loaded onto a 300-cycle NextSeq 500/550 HighOutput Kit v2 and run on the Illumina NextSeq 550 instrument (Illumina, San Diego, CA, USA). For Oxford Nanopore sequencing, the Midnight primer kit was used as described by Freed and Silander55. cDNA synthesis was performed on the extracted RNA using LunaScript RT mastermix (New England BioLabs) followed by gene-specific multiplex PCR using the Midnight Primer pools which produce 1200bp amplicons which overlap to cover the 30-kb SARS-CoV-2 genome. Amplicons from each pool were pooled and used neat for barcoding with the Oxford Nanopore Rapid Barcoding kit as per the manufacturer’s protocol. Barcoded samples were pooled and bead-purified. After the bead clean-up, the library was loaded on a prepared R9.4.1 flow-cell. A GridION X5 or MinION sequencing run was initiated using MinKNOW software with the base-call setting switched off. We assembled paired-end and nanopore.fastq reads using Genome Detective 1.132 (https://www.genomedetective.com) which was updated for the accurate assembly and variant calling of tiled primer amplicon Illumina or Oxford Nanopore reads, and the Coronavirus Typing Tool. For Illumina assembly, GATK HaploTypeCaller -- min-pruning 0 argument was added to increase mutation calling sensitivity near sequencing gaps. For Nanopore, low coverage regions with poor alignment quality (<85% variant homogeneity) near sequencing/amplicon ends were masked to be robust against primer drop-out experienced in the Spike gene, and the sensitivity for detecting short inserts using a region-local global alignment of reads, was increased. In addition, we also used the wf_artic (ARTIC SARS-CoV-2) pipeline as built using the nextflow workflow framework. In some instances, mutations were confirmed visually with .bam files using Geneious software V2020.1.2 (Biomatters). The reference genome used throughout the assembly process was NC_045512.2 (numbering equivalent to MN908947.3). To determine which SARS-CoV-2 proteins were mutated, sequence was input into the sequence analysis application in the Stanford Coronavirus Antiviral and Resistance Database (https://covdb.stanford.edu/sierra/sars2/by-sequences/) with HTML as output. Mutations were then visualized in Excel relative to the infecting variant.

### Phylogenetic analysis

Sequences were aligned by Nextclade version 2.9.1 (https://clades.nextstrain.org/). The json file output from the Nextclade analysis was loaded into Auspice (https://auspice.us/). Visualization was filtered to include reference sequences from clades 20A, 20B, 20H (Beta), 21J (Delta), 21M, 21K, 21L and 22B (Omicron), and the input sequences (new nodes), for a combined 1408 genomes. The tree was then filtered to show new nodes only. Tip labels were removed and SVG downloaded for final processing using Microsoft Powerpoint software.

### Cells

The H1299-E3 (H1299-ACE2, clone E3) cell line used in the live virus infections was derived from H1299 (CRL-5803) as described in previous work^55,67^ and propagated in growth medium consisting of complete Roswell Park Memorial Institute (RPMI) 1640 with 10% fetal bovine serum containing 10mM of HEPES, 1mM sodium pyruvate, 2mM L-glutamine and 0.1mM nonessential amino acids.

### Live virus neutralization assay

H1299-E3 cells were plated in a 96-well plate (Corning) at 30,000 cells per well 1 day pre-infection. Plasma was separated from EDTA-anticoagulated blood by centrifugation at 500 rcf for 10 min and stored at −80 °C. Aliquots of plasma samples were heat-inactivated at 56 °C for 30 min and clarified by centrifugation at 10,000 rcf for 5 min. Virus stocks were used at approximately 50-100 focus-forming units per microwell and added to diluted plasma. Antibody–virus mixtures were incubated for 1 h at 37 °C, 5% CO_2_. Cells were infected with 100 μL of the virus–antibody mixtures for 1 h, then 100 μL of a 1X RPMI 1640 (Sigma-Aldrich, R6504), 1.5% carboxymethylcellulose (Sigma-Aldrich, C4888) overlay was added without removing the inoculum. Cells were fixed 18 h post-infection using 4% PFA (Sigma-Aldrich) for 20 min. Foci were stained with a rabbit anti-spike monoclonal antibody (BS-R2B12, GenScript A02058) at 0.5 μg/mL in a permeabilization buffer containing 0.1% saponin (Sigma-Aldrich), 0.1% BSA (Sigma-Aldrich) and 0.05% Tween-20 (Sigma-Aldrich) in PBS. Plates were incubated with primary antibody overnight at 4 °C, then washed with wash buffer containing 0.05% Tween-20 in PBS. Secondary goat anti-rabbit HRP conjugated antibody (Abcam ab205718) was added at 1 μg/mL and incubated for 2 h at room temperature with shaking. TrueBlue peroxidase substrate (SeraCare 5510-0030) was then added at 50 μL per well and incubated for 20 min at room temperature. Plates were imaged in an ImmunoSpot Ultra-V S6-02-6140 Analyzer ELISPOT instrument with BioSpot Professional built-in image analysis (C.T.L).

### Statistics and fitting

All statistics and fitting were performed using custom code in MATLAB v.2019b. Neutralization data were fit to:

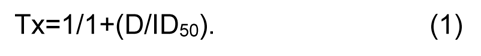

Here Tx is the number of foci at plasma dilution D normalized to the number of foci in the absence of plasma on the same plate. ID_50_ is the plasma dilution giving 50% neutralization.

FRNT_50_ = 1/ID_50_. Values of FRNT_50_ <1 are set to 1 (undiluted), the lowest measurable value. We note that the most concentrated plasma dilution was 1:25 and therefore FRNT_50_ < 25 were extrapolated.

### SARS-CoV-2 spike enzyme-linked immunosorbent assay (ELISA)

Two μg/mL of spike protein was used to coat 96-well, high-binding plates and incubated overnight at 4°C. The plates were incubated in a blocking buffer consisting of 5% skimmed milk powder, 0.05% Tween 20, 1x PBS. Plasma samples were diluted to 1:100 starting dilution in a blocking buffer and added to the plates. Secondary antibody was diluted to 1:3000 or 1:1000 respectively in blocking buffer and added to the plates followed by TMB substrate (Thermofisher Scientific). Upon stopping the reaction with 1 M H_2_SO_4_, absorbance was measured at a 450nm wavelength. MAbs CR3022 and BD23 were used as positive controls and Palivizumab was used as a negative control.

### SARS-CoV-2 hamster infections

Golden Syrian hamsters (4-5 weeks old) were purchased from Charles River Laboratories, USA. Experimental work was approved by the Animal Ethics Committee at the University of KwaZulu Natal (reference: REC/00004197/2022). For SARS-CoV-2 infection, hamsters were lightly sedated with 3% isoflurane (Piramal Healthcare, Mumbai, India) and infected with virus by intranasal inoculation of 50 µL per nostril of virus solution. Plasma of infected animals and uninfected controls was collected at 16 days post infection using cardiac puncture under anaesthesia with 5% isoflurane. Hamsters were immediately euthanized post-puncture with 1 mL of 200 mg/mL sodium pentobarbitone solution (Bayer AG, Leverkusen, Germany). Plasma was separated by centrifugation at 1000 x *g* for 10 min. Aliquots of plasma samples were heat-inactivated at 56 °C for 30 min and clarified by centrifugation at 10,000 rcf for 5 min.

## Data Availability

All data produced in the present work are contained in the manuscript

## Acknowledgements

This study was supported by the Bill and Melinda Gates award INV-018944 (AS), Wellcome Trust Award 226137/Z/22/Z (AS and PLM) and the South African Medical Research Council (AS and PLM). PLM is supported by the South African Research Chairs Initiative of the Department of Science and Innovation and National Research Foundation of South Africa. The funders had no role in study design, data collection and analysis, decision to publish, or preparation of the manuscript. AS wishes to thank Ron Milo, Karen Makar, and Thandi Onami for discussion and suggestions on analysis and presentation of results.

## Author contributions

AS and FK conceived the study and designed the study and experiments. FK, KK, MB, JU, GL, QAK, SSAK, and AvG identified and provided virus samples. M-YSM, FK, BIG, MB, KK, NM, NMa, MM, and NN set up and managed the cohort and cohort data. KK, ZJ, KR, YG, EV, ZM, and KG performed experiments and sequence analysis with input from AS, TdO, RJL. AS, FK, MB, JU, RJL, and GL interpreted data with input from M-YSM, SSAK, WH, and TdO. AS, FK, and GL prepared the manuscript with input from all authors.

---

**Figure S1:**
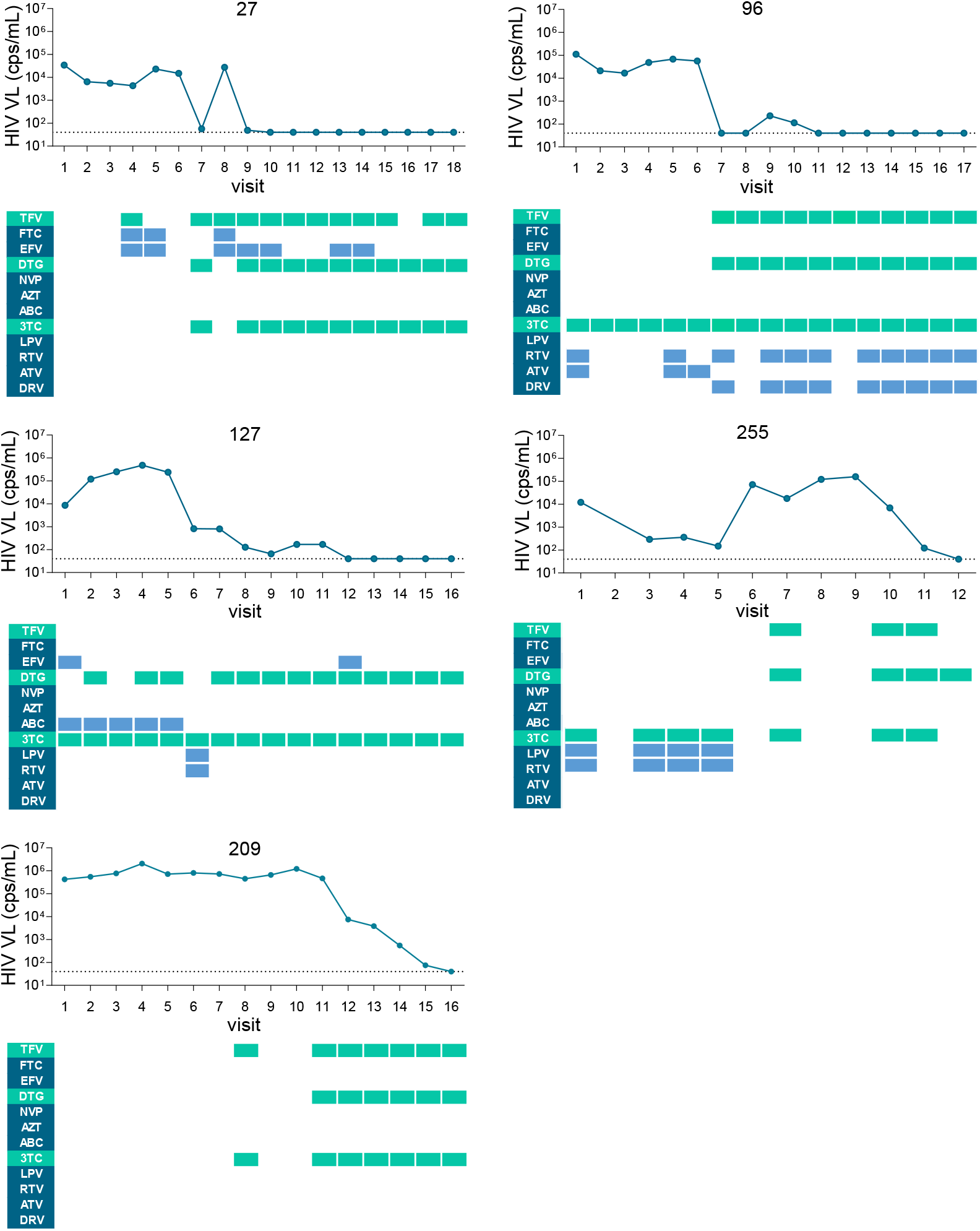
Antiretroviral drugs detected in plasma samples from the 5 participants with advanced HIV disease. ART components assayed by LC-MS/MS were the integrase inhibitor dolutegravir (DTG), the nucleotide reverse transcriptase inhibitor tenofovir (TFV), the nucleoside reverse transcriptase inhibitors emtricitabine (FTC), lamivudine (3TC), abacavir (ABC), and azidothymidine (AZT), the nonnucleoside reverse transcriptase inhibitors Efavirenz (EFV) and nevirapine (NVP), and the protease inhibitors lopinavir (LPV), ritonavir (RTV), atazanavir (ATV) and darunavir (DRV). X-axis is participant study visit and y-axis is HIV viral load as RNA copies/mL (above) or antiretroviral drug (below). Green or blue rectangles indicate the corresponding drug was detected at the timepoint at a level above 3 ng/mL, the limit of detection. Green rectangles indicate the presence of components of the regimen of TFV, 3TC, and DTG on which participants were initiated, and blue rectangles indicate components of other regimens.

**Figure S2:**
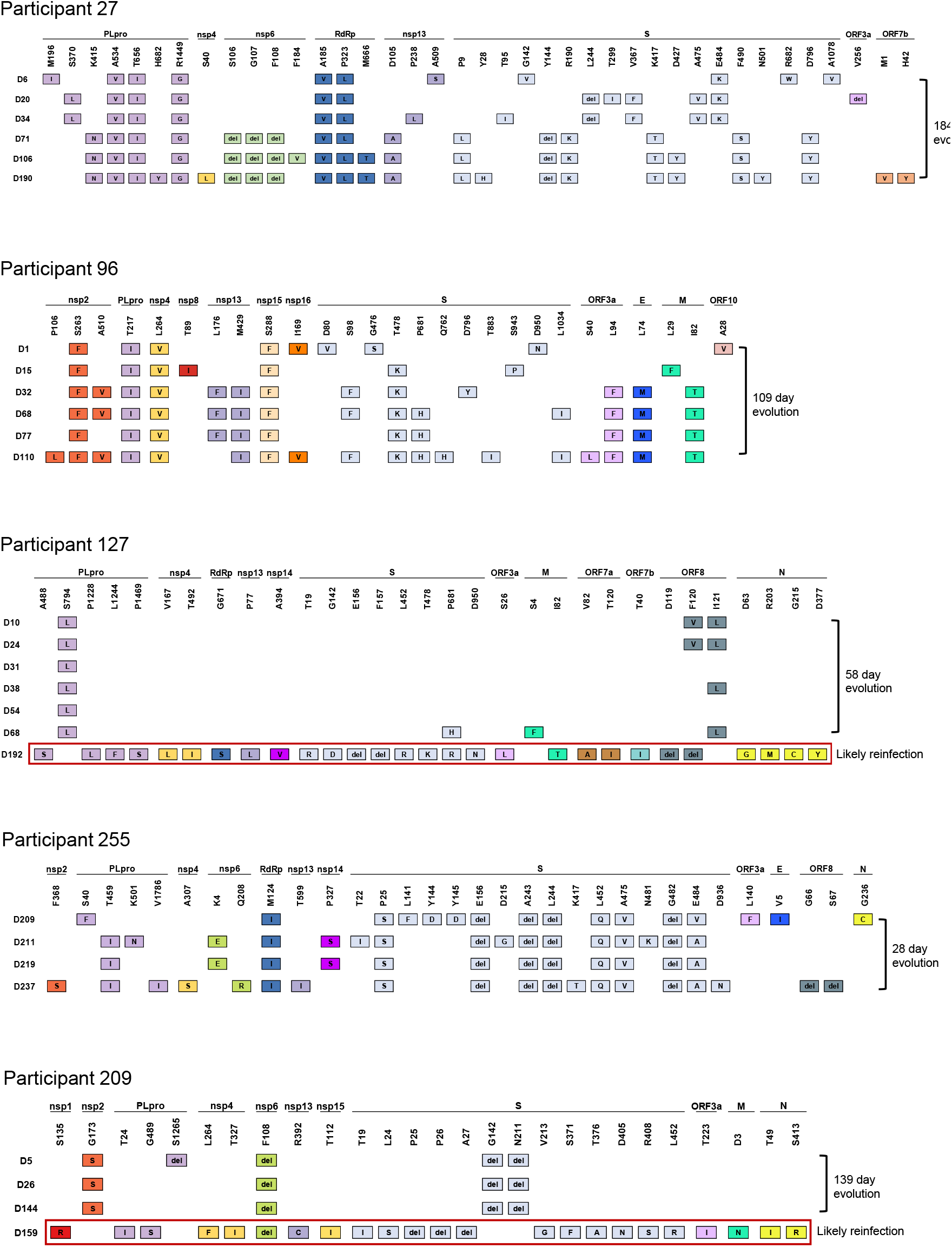
Substitutions or deletions in SARS-CoV-2 sequences of advanced HIV disease participants through time. Horizontal axis indicates the SARS-CoV-2 protein where substitution or deletion occurred relative to the infecting strain and vertical axis is the time post-diagnosis the viral isolate was obtained. Analysis performed using the Stanford Coronavirus Antiviral and Resistance Database (https://covdb.stanford.edu).

**Figure S3:**
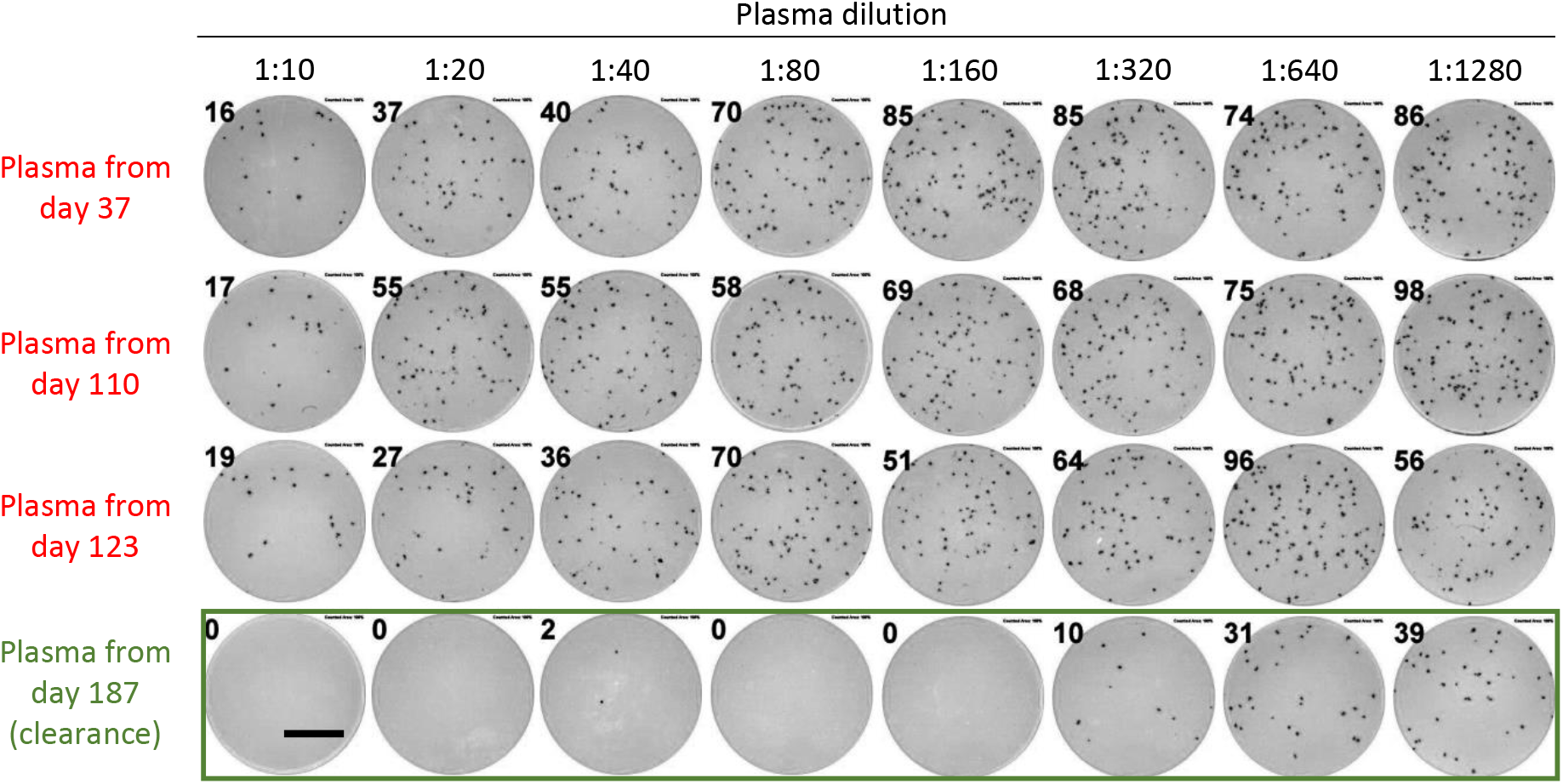
Neutralizing antibody response of participant plasma against autologous outgrown virus at different timepoints. Representative image showing infection foci in wells of a multi-well plate from a live virus focus forming assay of participant 209 plasma against autologous virus (209-D5). Plasma samples were from timepoints pre-SARS-CoV-2 clearance (D37, D110, D123) through to clearance (D187). Columns are plasma dilutions which range from 1:10 to 1:1280 and rows are plasma timepoint used. Bar is 2mm.

**Table S1:** Participants enrolled in the study.

|  | All<br>994 | HIV-<br>n=587 (59%) | HIV<br>Suppressed*<br>CD4** >200<br>n=259 (26%) | HIV viremic<br>CD4** >200<br>n=35 (4%) | HIV<br>viremic/suppressed<br>CD4** <200<br>n=113 (11%) |
| --- | --- | --- | --- | --- | --- |
| **Age (median, IQR)** | 41 (32-52) | 42 (31-55) | 42 (35-50) | 34.5 (27-38) | 41 (32-47) |
| **Female** | 621 (62%) | 347 (59%) | 193 (75%) | 22 (63%) | 59 (52%) |
| **SuppO2** | 143 (14%) | 62 (11%) | 36 (14%) | 8 (23%) | 37 (33%) |
| **CD4 count (median, IQR)** | 774.5 (467-1035) | 907.5 (676-1149) | 695 (478-901.5) | 433 (283-540) | 65 (23-134) |
| **Vaccination: Current Status of Study** |  |  |  |  |  |
| **Not vaccinated** | 493 (50%) | 253 (43%) | 127 (49%) | 24 (69%) | 89 (79%) |
| **1 dose BNT162b2\*\*\*** | 32 (3%) | 18 (3%) | 10 (4%) | 1 (3%) | 3 (3%) |
| **1 dose Ad26.COV2.S** | 281 (28%) | 210 (36%) | 59 (23%) | 6 (17%) | 6 (5%) |
| **2 doses BNT162b2** | 188 (19%) | 106 (18%) | 63 (24%) | 4 (11%) | 15 (13%) |
| **Variant#:** |  |  |  |  |  |
| **Ancestral** | 147 (15%) | 86 (14%) | 44 (17%) | 8 (23%) | 9 (8%) |
| **Beta** | 135 (13%) | 92 (16%) | 24 (9%) | 4 (11%) | 15 (13%) |
| **Delta** | 106 (11%) | 52 (9%) | 32 (12%) | 3 (9%) | 19 (16%) |
| **Omicron BA.1, BA.2** | 97 (10%) | 51 (9%) | 23 (9%) | 3 (9%) | 20 (18%) |
| **Omicron BA.5 related** | 53 (5%) | 22 (4%) | 14 (6%) | 4 (11%) | 13 (12%) |
| **Omicron XBB** | 96 (10%) | 65 (11%) | 21 (8%) | 2 (6%) | 8 (7%) |
| **No infection (controls)** | 360 (36%) | 219 (37%) | 101 (39%) | 11 (31%) | 29 (26%) |
| **Comorbidities:** |  |  |  |  |  |
| **Diabetes** | 108 (11%) | 77 (13%) | 26 (10%) | 1 (3%) | 4 (4%) |
| **Hypertension** | 199 (20%) | 129 (22%) | 57 (22%) | 4 (11%) | 9 (8%) |
| **Cardiovascular Disease** | 30 (3%) | 25 (4%) | 3 (1%) | 0 (0%) | 2 (2%) |

\*Median HIV viral load < 500. \*\*Median CD4 count. \*\*\*At enrollment. #Variant determined by dominant circulating variant at enrollment.

**Table S2:** Participants with advanced HIV disease in the study.

| Participant | Sex | Age range | Diagnosis | Infect. wave | Enrol. CD4 | Enrol. HIV VL | 1<sup>st</sup> Vacc. Date | Vacc. CD4 | Vacc. HIV VL | Time to last positive |
| --- | --- | --- | --- | --- | --- | --- | --- | --- | --- | --- |
| **27** | F | 30-39 | Sep 2020 | D614G | 6 | 34151 | Sep 2021 | 92 | 40 | 215 |
| **96** | M | 40-49 | Apr 2021 | Beta | 4 | 111883 | Nov 2021 | 59 | 40 | 110 |
| **127** | M | 30-39 | Mar 2021 | Beta | 12 | 8581 | Sep 2021 | 120 | 40 | 207 |
| **209** | F | 30-29 | Dec 2021 | Omicron | 24 | 423817 | Apr 2022 | 31 | 661666 | 165 |
| **255** | M | 20-29 | Sep 2021 | Delta | 2 | 12041 | Jun-2022 | 42 | 17813 | 289 |

**Table S3:**
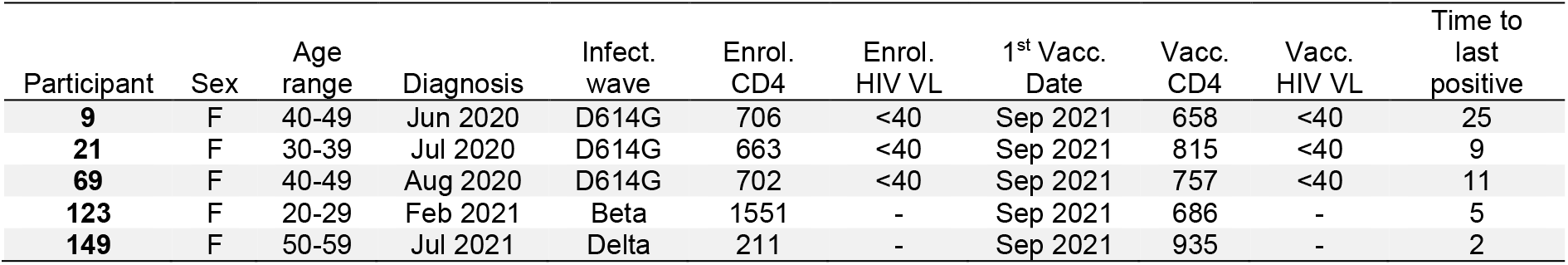
Pfizer-vaccinated longitudinally tracked participant controls.

**Table S4:**
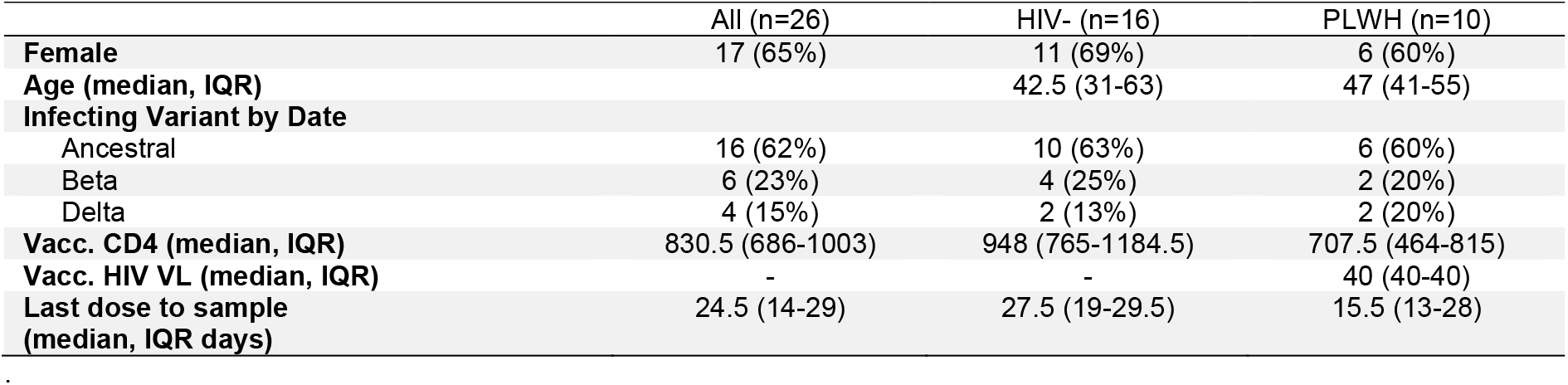
Summary vaccinated participants (no advanced HIV disease)

**Table S5:**
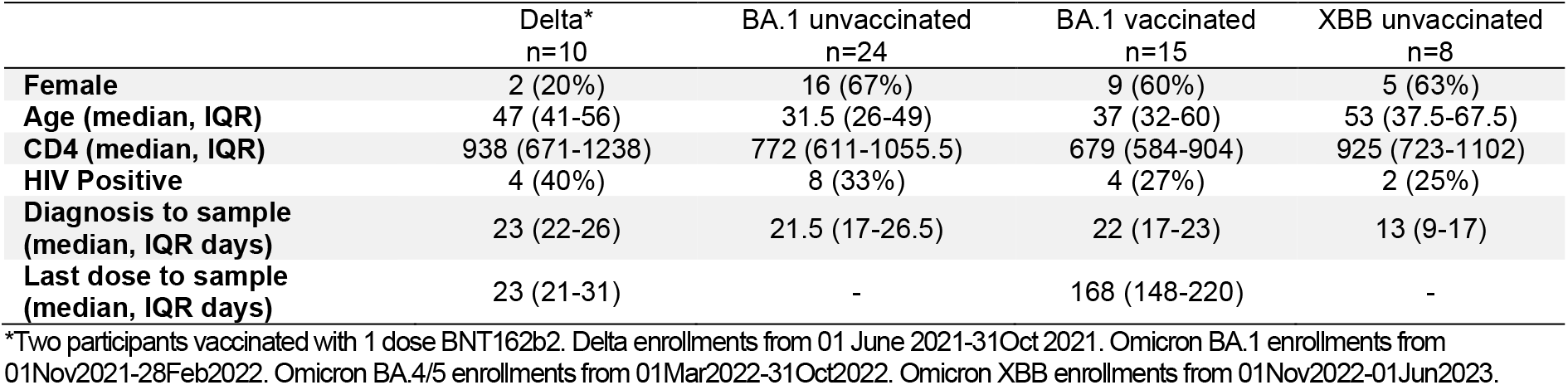
Participants from different infection periods.

|  | Delta*<br>n=10 | BA.1 unvaccinated<br>n=24 | BA.1 vaccinated<br>n=15 | XBB unvaccinated<br>n=8 |
| --- | --- | --- | --- | --- |
| **Female** | 2 (20%) | 16 (67%) | 9 (60%) | 5 (63%) |
| **Age (median, IQR)** | 47 (41-56) | 31.5 (26-49) | 37 (32-60) | 53 (37.5-67.5) |
| **CD4 (median, IQR)** | 938 (671-1238) | 772 (611-1055.5) | 679 (584-904) | 925 (723-1102) |
| **HIV Positive** | 4 (40%) | 8 (33%) | 4 (27%) | 2 (25%) |
| **Diagnosis to sample (median, IQR days)** | 23 (22-26) | 21.5 (17-26.5) | 22 (17-23) | 13 (9-17) |
| **Last dose to sample (median, IQR days)** | 23 (21-31) | - | 168 (148-220) | - |

\*Two participants vaccinated with 1 dose BNT162b2. Delta enrollments from 01 June 2021-31Oct 2021. Omicron BA.1 enrollments from 01Nov2021-28Feb2022. Omicron BA.4/5 enrollments from 01Mar2022-31Oct2022. Omicron XBB enrollments from 01Nov2022-01Jun2023.

